# Calibrating self-reported BMI in national surveillance: impact on obesity misclassification and socioeconomic inequalities in Portugal

**DOI:** 10.64898/2026.08.24.26357362

**Authors:** Berta Valente, Catarina Campos Silva, Milton Severo, Andreia Oliveira, Ulf-Göran Gerdtham, Joana Araújo

## Abstract

**Background:** Self-reported height and weight are prone to misreporting, which can bias BMI estimates. This study identifies misreporting determinants, develops calibration equations and examines how measured, self-reported, and calibrated BMI affect estimates of obesity prevalence and socioeconomic inequalities.

**Methods:** We analysed survey-weighted, sex stratified data from 3,404 adults (18-64 years) in the Portuguese National Food, Nutrition and Physical Activity Survey (IAN-AF 2015–2016), including self-reported and measured anthropometry. Misreporting determinants were assessed using multinomial logistic regression. Calibration equations for height and weight were estimated using measured values, self-reports, age, region of residence and education level. Calibrated BMI was derived from predicted values. Obesity prevalence was estimated for each BMI assessment method (≥ 30 *kg/m*^2^). Education, income and employment inequalities in obesity were compared across BMI methods using prevalence difference and ratio, slope index and relative indexes of inequality.

**Results:** Height is systematically overreported and weight underreported, with misreporting increasing with age and BMI. Calibration eliminates underestimation of obesity prevalence from self-reported BMI, bringing calibrated estimates close to measured values. Regarding education-related inequalities in obesity, calibration widen disparities among women, whereas among men corrects the overestimation observed from self-reported BMI. Income and employment-inequality patterns are similar across BMI methods.

**Conclusions:** Among Portuguese adults, the systematic and socially patterned misreport of self-reported anthropometry affects obesity prevalence and inequality estimates. Calibration based on simple sociodemographic models improves validity and equity of obesity surveillance and could be routinely integrated into national surveys to strengthen monitoring of obesity and its socioeconomic distribution.

**Key points:**

- The magnitude and determinants of misreporting in height and weight vary by sex and socioeconomic characteristics and are context-specific, indicating that misreporting is not uniform across population groups or settings.
- Simple sex-specific calibration equations using routinely collected variables (age, sex, region, education) substantially reduce obesity misclassification.
- Calibrated BMI almost fully aligns obesity prevalence with measured BMI, correcting underestimation from self-reported data in both sexes.
- Calibration changes education-related obesity inequalities, slightly widening them among women and attenuating their overestimation among men.
- Wider adoption of similar calibration approaches in European national surveys that rely on self-reported BMI could improve the validity of obesity surveillance and inequality monitoring.

## Introduction

National health surveys and large epidemiological studies are cemtral to monitoring estimates of obesity prevalence, trends and socioeconomic inequalities and to informing public health policy (1), but many rely on self-reported anthropometry due to time and budget constrains (2). Self-reports are prone to misreporting, and beyond recall and rounding, the magnitude and direction of error vary systematically by demographic, socioeconomic characteristics and tend towards socially desirable responses (3–6). In general, men tend to overreport height and women to underreport weight (7), resulting in body mass index (BMI) underestimation of up to two units in some settings (8,9). Additionally, self-reported BMI typically overestimates lower values and underestimates higher values relative to measured BMI (10). Because many national and international analyses rely on these self-reported data, and misreporting is socially patterned, such bias can complicate comparisons of obesity levels and inequalities across populations and over time.

Such misreporting can induce systematic BMI misclassification (11), bias estimates when BMI is used as an outcome or exposure (12), and distort estimates of education- and income-related inequalities in obesity (12). One approach to mitigate these biases is to apply correction factors or calibration equations to self-reported data, often incorporating sociodemographic predictors to improve the accuracy of BMI estimates (2,10,13–17). Calibration equations can improve agreement between self-reported and measured BMI, but they require both self-reported and objectively measured anthropometry. They are also typically context-specific, hindering generalisability and broader application (2).

In Portugal, documented socioeconomic-related inequalities in obesity rely on self-reported anthropometric data(18). Therefore, this evidence is susceptible to systematic reporting bias, as even modest differential misreporting across socioeconomic position (SEP) may bias inequality estimates (19). The Portuguese National Food, Nutrition and Physical Activity Survey (IAN-AF 2015-2016) contains both measured and self-reported data (20), providing an ideal setting to evaluate whether calibration improves obesity prevalence estimates and socioeconomic inequalities. While prior Portuguese work has examined the validity of self-reported anthropometry (21) and described temporal trends in obesity inequalities (18), none have combined concurrent measured and self-reported data in a nationally representative survey to derive calibration equations and assess their impact on inequality estimates across BMI measures. We therefore aim to: (1) identify demographic and socioeconomic determinants of misreporting in self-reported weight and height; (2) develop and internally validate sex-specific calibration equations based on routinely collected survey variables; and (3) assess how measured, self-reported, and calibrated BMI affect obesity misclassification and associated socioeconomic inequalities.

## Methods

### Study Participants

This cross-sectional study uses data from IAN-AF (October 2015-September 2016), a nationally representative survey of the Portuguese population aged 3 months to 84 years, based on a multistage, stratified sampling design (20). In the first sampling stage, primary health care units were randomly selected within the seven Statistical Geographic Units of Portugal (NUTS II) and weighted by the number of individuals registered in each unit (20). In the second stage, individuals were randomly selected from these registries, stratified by sex and age group (20). Data were collected by trained fieldworkers using computer-assisted personal interviewing, with two face-to-face interviews conducted at home or in a primary health care setting, according to participant preference (20). Of the 6,553 individuals completing the first interview, 3404 (51.9%) adults aged 18-84 years with complete self-reported and measured anthropometry were included in the present analysis. Exclusions are shown in Figure S1. Compared with those not included, included participants were more likely to have higher education (25.0% vs 14.8%) and to be employed (59.9% vs 40.1%) (Supplementary Table S1).

### Anthropometric Measurements

Self-reported height and weight were collected before objective assessment, followed by measurements by trained fieldworkers using standardised procedures (22). BMI was calculated and analysed continuously for bias and agreement. For misclassification and inequality analyses, BMI was categorised as underweight (<18.5 kg/m²), normal weight (18.5– 24.9 kg/m²), pre-obesity (25.0–29.9 kg/m²) and obesity (≥30.0 kg/m²) (23), with underweight combined with normal weight due to low prevalence (0.9%). The main outcome is BMI assessed from measured, self-reported, and calibrated data.

### Demographic, socioeconomic and health-related characteristics

Demographic characteristics include age (years), sex (women/men) and region (North, Centre, Lisbon Metropolitan Area, Alentejo, Algarve, and the Autonomous Regions of Madeira and the Azores). Socioeconomic characteristics include: education, the highest completed level of schooling, grouped as low (up to 9th grade), medium (12th grade or post-secondary), or high (bachelor’s, master’s, or doctoral degree); income, the net monthly household income after deductions for taxes and social, classified as low (<€970), medium (€971–1,940), or high (>€1,940); and employment status, categorised as employed (for pay or profit, including unpaid work in a family business or farm, apprenticeship or paid internship, and workers on maternity, paternity, sick, or vacation leave), unemployed (without a job during the reference period, available for work, and actively seeking employment), or other (retired, permanently disabled, student, household worker, or performing compulsory military or community service) (20).

### Measures of inequality

Inequality is assessed using simple measures, prevalence differences (PD) and prevalence ratios (PR), comparing extreme categories of socioeconomic position (high *versus* low, with the highest category as reference); and regression-based measures, the slope index of inequality (SII) and the relative index of inequality (RII), which summarise absolute and relative inequalities across the full socioeconomic distribution by ranking socioeconomic categories and modelling them as RIDIT scores (24).

### Statistical Analysis

All analyses account for complex survey design (strata, primary sampling units, and sampling weights), ensuring national representativeness, and were stratified by sex. Sex stratification was decided a priori, given known sex differences in misreporting and obesity inequalities (12). Analyses were guided by a conceptual framework for systematic, socially patterned misreporting and its implications for inequality measures (Supplementary Material, Section S1).

Analyses were conducted using complete cases. Income had approximately 10% missing values, whereas all other covariates <3%; missingness patterns and differences between complete and incomplete cases are shown in Supplementary Table S1. Robustness of the results was assessed by a sensitivity analysis using multiple imputation by chained equations (MICE; 20 imputed datasets), including all analysis variables in the imputation model and combined estimates using Rubin’s rules (25) (Supplementary Table S10). Unweighted participant characteristics are summarised in Supplementary Table S2. Agreement between measured and self-reported height/weight were examined using Bland-Altman plots (Figure S1). Differences between self-reported and measured height/weight (reporting bias) across sociodemographic characteristics were estimated using survey-adjusted linear regression (Supplementary Tables S3-S4). Age-related patterns of reporting bias in height, weight and BMI were explored using LOESS (95% confidence intervals (CI)), stratified by measured BMI (Figures S2-S3).

Prior Portuguese validation considered differences within ±0.5 cm for height and ±1 kg for weight as no bias (21), thus misreporting determinants were examined using survey-weighted multinomial logistic regression, with under- and overreporting *versus* no bias as the outcome (Odds Ratio (OR), 95%CI). No bias was considered for height and weight differences within ±0.5 cm and ±1 kg, respectively, according to previous literature (21). Models were adjusted for age, region, education, income, employment status, and measured BMI. Region was modelled using effects (sum-to-zero) coding, so that estimates reflect deviations from the national mean (26). As sensitivity analyses, we tested a wider threshold to define no bias (height: ±1.0 cm, weight: ±2.0 kg; Supplementary Table S3), and examined the absolute magnitude of reporting bias using survey-adjusted linear regression (Supplementary Tables S4-S5).

To address reporting bias, we developed sex-specific indirect calibration models by regressing measured height/weight on self-reported values, age, region, and education. For height, the model was:

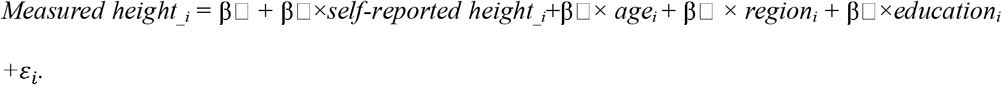

and analogously for weight. Estimated coefficients and corresponding 95%CIs are presented in Supplementary Table S6. Backward variable selection was used to obtain parsimonious models, excluding income and employment status, as simpler models may outperform more complex specifications (27). Calibrated BMI was computed from calibrated height and weight. Estimated coefficients and worked examples illustrating the application of the calibration equations are provided in Box S1. Calibration equations were internally validated using a 70/30 split□sample design, with sex□specific models re□estimated in the training set and assessed in the test set by calibration□in□the□large (measured BMI-calibrated BMI) and calibration slope (measured regressed on calibrated BMI).

Survey-weighted means and standard errors (SE) for measured, self-reported, and calibrated anthropometrics are reported in Supplementary Table S7. Agreement between BMI assessment methods was investigated using a survey-weighted approximation of Lin’s concordance correlation coefficient (CCC) and agreement between calibrated and measured BMI was defined as the percentage classified into the same category (Supplementary Tables S8–S9). Prevalence estimates for each BMI assessment method are obtained as survey-weighted proportions. Sex-specific socioeconomic inequalities in obesity were assessed using PD, PR, SII and RII along with 95% CIs, and stratified by BMI assessment method. Analyses were performed in R (v.4.4.8) with survey, mice and VGAM packages (28).

### Ethics

Ethical approval was obtained from the National Commission for Data Protection (4940/2015 from 26th May, 2015), the Ethical Committee of the Institute of Public Health of the University of Porto (CE15033, from 13th March, 2015) and the Ethical Commissions of each of the Regional Administration of Health. All participants were also asked to provide their written informed consent for participation according to the Ethical Principles for Medical Research involving human subjects expressed in the Declaration of Helsinki and the national legislation.

## Results

### Study population

The analytical sample represents approximately 7.2 million Portuguese adults (48.0% women). Table 1 describes participant’s characteristics.

**Table 1.** Weighted characteristics of Portuguese adults by sex, IAN-AF 2015–2016.

|  | **Total, (%)**<br>7,214,614 |  |
| --- | --- | --- |
|  | **Women**<br>3,461,077 (48.0) | **Men**<br>3,753,537 (52.0) |
| **Age, mean (SD)** | 46.7 (16.0) | 46.9 (16.5) |
| **Region** |  |  |
| North | 1,289,134 (37.2) | 1,444,260 (38.5) |
| Center | 737,955 (21.3) | 769,270 (20.5) |
| Lisbon Metropolitan Area | 882,529 (25.5) | 941,159 (25.1) |
| Alentejo | 209,501 (6.1) | 240,081 (6.4) |
| Algarve | 156,421 (4.5) | 161,850 (4.3) |
| Autonomous Region of Madeira | 95,476 (2.8) | 101,710 (2.7) |
| Autonomous Region of the Azores | 90,061 (2.6) | 95,208 (2.5) |
| **Education level** |  |  |
| Low | 1,559,976 (45.2) | 1,899,588 (50.7) |
| Intermediate | 985,605 (28.5) | 1,025,222 (27.3) |
| High | 907,152 (26.3) | 823,760 (22.0) |
| **Employment Status** |  |  |
| Employed | 1,939,611 (56.3) | 2,193,384 (58.4) |
| Other employment situation | 1,092,707 (31.7) | 1,082,289 (28.8) |
| Unemployed | 413,702 (12.0) | 477,865 (12.7) |
| **Income** |  |  |
| Low | 1,176,407 (38.6) | 1,119,455 (32.4) |
| Intermediate | 1,225,129 (40.2) | 1,403,484 (40.7) |
| High | 647,568 (21.2) | 926,911 (26.9) |
| **Weight (kg), mean (SD)** |  |  |
| Self-reported | 66.0 (11.3) | 78.3 (12.0) |
| Measured | 66.7 (11.8) | 78.9 (12.9) |
| **Height (cm), mean (SD)** |  |  |
| Self-reported | 160.3 (6.2) | 172.7 (7.0) |
| Measured | 158.2 (6.1) | 171.6 (7.0) |

Abbreviations: SD, standard deviation

### Patterns of reporting bias

Systematic reporting bias is observed, with weight generally underreported and height overreported. Discrepancies are larger at higher measured values (Figure S2, Figure S3). The magnitude of bias increases with age in both sexes (Figure S4). Among women, height overreporting and weight underreporting are more pronounced among those with obesity, whereas among men, BMI-related differences are more evident for weight underreporting (Figure S3).

### Determinants of height and weight misreporting

Misreporting determinants are shown in Table 2. Among women, height underreporting is more frequent in Madeira (OR:2.17, 95%CI: 1.22–3.86) and Azores (1.54, 1.05–2.24), while height overreporting is more frequent in the North (1.88, 1.30–2.70) and less frequent in Algarve (0.58, 0.42-0.81). Weight underreporting increased with BMI, with women with pre-obesity and obesity having approximately 1.6-fold (1.57, 1.05–2.33) and 2.8-fold (2.76, 1.67–4.55) higher odds of underreporting, respectively. Regional variation in weight underreporting was modest, with higher odds in the Center (1.51, 1.14-2.00) and lower odds in Alentejo (0.69, 0.49–0.97).

**Table 2.** Determinants of <u>height</u> (± 0.5 cm) and <u>weight</u> (± 1 kg) bias relative to measured values, stratified by sex.

|  | Height |  |  |  | Weight |  |  |  |
| --- | --- | --- | --- | --- | --- | --- | --- | --- |
|  | Women |  | Men |  | Women |  | Men |  |
|  | OR (95% CI) |  | OR (95% CI) |  | OR (95% CI) |  | OR (95% CI) |  |
|  | *Underreport vs no bias* | *Overreport vs no bias* | *Underreport vs no bias* | *Overreport vs no bias* | *Underreport vs no bias* | *Overreport vs no bias* | *Underreport vs no bias* | *Overreport vs no bias* |
| Age (years) | 1.03 (1.01–1.05) | 1.03 (1.01–1.04) | 0.99 (0.97–1.01) | 0.99 (0.98–1.01) | 1.01 (0.99–1.02) | 1 (0.99–1.02) | 1.01 (1–1.03) | 0.99 (0.98–1.01) |
| **Region** |  |  |  |  |  |  |  |  |
| North | 1.5 (0.9–2.51) | **1.88 (1.3–2.7)** | 1.42 (0.99–2.04) | 1.1 (0.81–1.5) | 0.93 (0.76–1.14) | 0.9 (0.61–1.33) | 1.04 (0.77–1.41) | 0.73 (0.49–1.09) |
| Center | 0.78 (0.48–1.27) | 1.00 (0.71–1.4) | 0.84 (0.6–1.17) | 1.04 (0.73–1.48) | **1.51 (1.14–2)** | 1.53 (0.98–2.39) | 0.89 (0.66–1.2) | 1.18 (0.83–1.7) |
| Lisbon M.A. | 0.58 (0.28–1.24) | 0.82 (0.54–1.23) | 0.78 (0.48–1.26) | 1.13 (0.74–1.74) | 1.01 (0.72–1.43) | 0.68 (0.39–1.16) | 0.96 (0.67–1.37) | 0.96 (0.67–1.37) |
| Alentejo | 0.57 (0.23–1.42) | 1.25 (0.72–2.14) | 1 (0.61–1.62) | 1.24 (0.93–1.66) | **0.69 (0.49–0.97)** | 1.16 (0.76–1.78) | 0.61 (0.38–0.97) | 1.54 (1.14–2.09) |
| Algarve | 0.77 (0.42–1.44) | **0.58 (0.42–0.81)** | 0.92 (0.64–1.32) | 0.78 (0.61–1.01) | 0.83 (0.61–1.12) | 1.34 (0.86–2.1) | 1.29 (0.95–1.77) | 0.64 (0.47–0.87) |
| A. R. Madeira | **2.17 (1.22–3.86)** | 1.13 (0.76–1.66) | 1.17 (0.92–1.49) | 0.87 (0.63–1.22) | 1.06 (0.86–1.3) | 0.72 (0.49–1.05) | 1.13 (0.87–1.46) | **1.38 (1.05–1.81)** |
| A.R. Azores | **1.54 (1.05–2.24)** | 0.8 (0.54–1.19) | 1.01 (0.73–1.39) | 0.9 (0.67–1.22) | 1.15 (0.94–1.4) | 0.96 (0.57–1.63) | 1.27 (0.94–1.73) | 0.89 (0.61–1.32) |
| **Education level** |  |  |  |  |  |  |  |  |
| Low | 0.65 (0.24–1.74) | 0.77 (0.41–1.44) | 1.26 (0.55–2.86) | 1.1 (0.57–2.13) | 0.84 (0.54–1.3) | 0.93 (0.53–1.62) | 0.51 (0.28–0.93) | 1.3 (0.69–2.46) |
| Intermediate | 1.22 (0.59–2.48) | 1.36 (0.79–2.32) | 1.19 (0.62–2.26) | 1.06 (0.6–1.88) | 1.13 (0.74–1.73) | 0.86 (0.48–1.52) | 0.68 (0.37–1.24) | 1.4 (0.79–2.48) |
| High | Ref. | Ref. | Ref. | Ref. | Ref. | Ref. | Ref. | Ref. |
| **Income level** |  |  |  |  |  |  |  |  |
| Low | 2.25 (0.84–6.06) | 1.07 (0.65–1.74) | 1.85 (0.84–4.07) | 0.91 (0.5–1.66) | 1.13 (0.7–1.82) | 1.7 (0.85–3.39) | 1.44 (0.87–2.39) | **2.6 (1.39–4.86)** |
| Intermediate | 2.15 (1.01–4.6) | 1.2 (0.77–1.9) | 0.97 (0.49–1.9) | 0.73 (0.43–1.26) | 0.81 (0.51–1.3) | 1.44 (0.77–2.7) | 1.16 (0.74–1.83) | **1.83 (1.12–3)** |
| High | Ref. | Ref. | Ref. | Ref. | Ref. | Ref. | Ref. | Ref. |
| **Employment status** |  |  |  |  |  |  |  |  |
| Employed | Ref. | Ref. | Ref. | Ref. | Ref. | Ref. | Ref. | Ref. |
| Other | 1.21 (0.61–2.41) | 0.85 (0.53–1.36) | 0.87 (0.46–1.68) | 1.43 (0.9–2.27) | 0.92 (0.59–1.43) | 0.75 (0.49–1.15) | 1.37 (0.83–2.25) | 0.85 (0.47–1.53) |
| Unemployed | 1.93 (0.79–4.71) | 0.87 (0.52–1.45) | 0.68 (0.27–1.73) | 0.89 (0.44–1.79) | 1.31 (0.79–2.19) | 1.18 (0.64–2.18) | 1.6 (0.91–2.8) | 1.03 (0.52–2.05) |
| **BMI category** |  |  |  |  |  |  |  |  |
| Normal | Ref. | Ref. | Ref. | Ref. | Ref. | Ref. | Ref. | Ref. |
| Pre-obesity | 1.54 (0.76–3.11) | 1.37 (0.88–2.14) | 0.99 (0.64–1.54) | 1.7 (1.14–2.53) | **1.57 (1.05–2.33)** | 0.45 (0.26–0.78) | **2.26 (1.51–3.37)** | 0.76 (0.48–1.21) |
| Obesity | 0.55 (0.23–1.31) | 2.03 (1.21–3.4) | 1 (0.52–1.92) | 1.1 (0.64–1.89) | **2.76 (1.67–4.55)** | 0.7 (0.35–1.38) | **4.64 (2.44–8.82)** | 0.76 (0.41–1.42) |

Notes: Adjusted odds ratios (ORs) were estimated using sex-stratified, survey-weighted multinomial logistic regression models. ORs greater than 1 indicate higher odds of under-reporting or over-reporting (as specified in the column heading) relative to no reporting bias. Regional ORs are expressed relative to the overall mean across all regions (effects coding). No reporting bias was defined as a difference between self-reported and measured height within $\pm 0.5$ cm, and between self-reported and measured weight within $\pm 1$ kg. Reporting bias categories were treated as nominal outcomes, with “no bias” set as the reference. Because effects coding estimates coefficients for $k - 1$ regions relative to the overall mean, OR (95% CI) for Azores region were derived from the remaining coefficients and their variance-covariance matrix.

Among men, height overreporting is more frequent among those with pre-obesity (1.70, 1.14-2.53). Weight underreporting is less common among men with low education (0.51, 0.28– 0.93) and in Algarve (0.61, 0.38–0.97), but more frequent among those with pre-obesity (2.26, 1.51–3.37) and obesity (4.64, 2.44–8.82). Weight overreporting is more frequent among men with low (2.60, 1.39–4.86) or intermediate income (1.83, 1.12–3.00) and those living in Madeira (1.38, 1.05-1.81). Results were similar using wider thresholds (±1.0 cm for height and ±2.0 kg for weight), with positive associations between misreporting and higher BMI and with education, income and region gradiens mantained (Supplementary Table S3).

### Calibration of self-reported height and weight

Calibration reduces discrepancies between self-reported and measured anthropometry (Supplementary Table S7), increases concordance between BMI assessment methods and improves agreement in BMI categories, with obesity agreement ≥86% in both sexes (Supplementary Tables S8–S9). Obesity prevalence from calibrated BMI more closely matches that from measured BMI among women (26.0% vs 26.2%) and men (21.7% vs 22.5%), correcting the underestimation observed with self-reported BMI (women: 25.3%, men: 20.0%; Table 3).

**Table 3.** Prevalence of BMI categories by BMI assessment method and sex.

|  |  | BMI measure |  |  |
| --- | --- | --- | --- | --- |
|  |  | Measured (%) | Self-reported (%) | Calibrated (%) |
| Women | Normal weight | 42.3 | 42.2 | 39.8 |
|  | Pre-obesity | 31.5 | 32.6 | 34.2 |
|  | Obesity | 26.2 | 25.3 | 26.0 |
| Men | Normal weight | 34.9 | 37.0 | 35.9 |
|  | Pre-obesity | 43.4 | 43.0 | 41.7 |
|  | Obesity | 21.7 | 20.0 | 22.5 |

Abbreviations: BMI, body mass index

### Impact of BMI measures on socioeconomic inequalities in obesity

Across all BMI measures, education-related inequalities in obesity are more pronounced, followed by income- and employment-related inequalities (Table 4). Among women, educational inequalities in obesity are similar across BMI assessment methods, with calibrated BMI yielding higher point estimates than measured and self-reported BMI. Among men, calibration mainly reduces the overestimation of education-related inequalities observed with self-reported BMI, bringing estimates closer to those based on measured BMI. Income-related inequalities are broadly similar across BMI methods in women, while in men calibrated BMI yields higher inequality estimates than measured BMI. For employment, calibrated BMI generally produces higher PD and PR estimates in both sexes; among men, SII and RII are lower with calibrated BMI than with measured BMI, whereas among women calibrated estimates are closer to those based on measured BMI. Sensitivity analyses using multiple imputation are consistent with complete-case findings, with inequality patterns across BMI methods largely unchanged (Supplementary Table S10).

**Table 4.** Socioeconomic inequalities in obesity by BMI assessment method, sex, and socioeconomic indicators (complete-case analysis)

|  |  | Education |  | Income |  | Occupation |  |
| --- | --- | --- | --- | --- | --- | --- | --- |
|  | BMI | PD | PR | PD | PR | PD | PR |
| Women | *Measured* | 23.0 (17–28.9) | 2.55 (1.92–3.39) | 11.9 (5.4–18.4) | 1.59 (1.23–2.06) | -4.5 (-14.9–5.9) | 0.82 (0.5–1.34) |
|  | *Self-reported* | 23.1 (17.7–28.6) | 2.83 (2.13–3.76) | 11.8 (5.3–18.4) | 1.66 (1.24–2.21) | -5.9 (-14.5–2.8) | 0.75 (0.46–1.21) |
|  | *Calibrated* | 25.7 (20.2–31.1) | 3.04 (2.32–3.98) | 11.7 (4.2–19.3) | 1.61 (1.19–2.19) | -2.2 (-11.2–6.7) | 0.91 (0.6–1.36) |
| Men | *Measured* | 9.2 (3.5–14.9) | 1.57 (1.17–2.11) | 8.5 (2.6–14.4) | 1.47 (1.12–1.94) | -4.8 (-13.6–4) | 0.77 (0.46–1.31) |
|  | *Self-reported* | 10.8 (5.7–15.9) | 1.92 (1.39–2.66) | 8.9 (3–14.7) | 1.62 (1.19–2.19) | -4.9 (-13.1–3.2) | 0.72 (0.39–1.34) |
|  | *Calibrated* | 8.6 (2.7–14.5) | 1.52 (1.13–2.04) | 11.8 (4.8–18.8) | 1.69 (1.25–2.28) | -8.3 (-16.8–0.2) | 0.62 (0.34–1.13) |
|  | BMI | SII | RII | SII | RII | SII | RII |
| Women | *Measured* | 42.0 (31.3–52.8) | 6.64 (3.54–12.46) | 25.6 (14–37.2) | 2.92 (1.71–5.01) | 19.4 (6.8–32) | 2.13 (1.34–3.38) |
|  | *Self-reported* | 42.2 (33–51.3) | 8.32 (4.71–14.7) | 25.9 (14.7–37.1) | 3.3 (1.86–5.84) | 15.5 (3.8–27.2) | 1.94 (1.2–3.13) |
|  | *Calibrated* | 46.4 (36.5–56.3) | 9.79 (5.43–17.65) | 25.6 (13.2–38.1) | 3.12 (1.73–5.62) | 18.9 (6.8–30.9) | 2.17 (1.36–3.47) |
| Men | *Measured* | 15.7 (5.3–26.2) | 2.22 (1.27–3.88) | 8.5 (-2.7–19.7) | 1.51 (0.86–2.65) | 3.9 (-7.1–14.8) | 1.2 (0.72–2) |
|  | *Self-reported* | 19.4 (10.9–27.9) | 3.39 (1.87–6.17) | 12.4 (2.9–21.9) | 2.07 (1.16–3.67) | 0.4 (-11.7–12.5) | 1.02 (0.51–2.06) |
|  | *Calibrated* | 14.4 (4.1–24.8) | 2.09 (1.19–3.67) | 14 (2.5–25.5) | 2 (1.12–3.58) | -1.3 (-13.9–11.4) | 0.94 (0.5–1.76) |

Abbreviations: PD, prevalence difference in percentual points; PR, prevalence ratio; SII, slope index of inequality; RII, relative index of inequality

## Discussion

This study examines patterns and determinants of misreporting in self-reported height and weight among Portuguese adults and how calibration influences estimates of obesity prevalence and socioeconomic inequalities. Self-reported anthropometry shows systematic and socially patterned misreporting, with height overreported, weight underreported, and bias increasing with age and BMI. Calibration improves agreement between self-reported and measured BMI, reduces obesity misclassification in both sexes, and alters socioeconomic inequality estimates: slighlty increasing education-related disparities in women, while attenuating overestimation in men.

The direction and magnitude of observed misreporting align with previous studies documenting height overestimation and weight underestimation across populations (4, 11, 24, 29). However, height overreporting in our study appears greater than in prior Portuguese work (21) and some international studies (4,11,29), which examined misreporting by sex, age, education, occupation, smoking and self-rated health (21) and showed that height overreporting and weight underreporting are more pronounced at older ages and higher BMI, leading to BMI underestimation and, consequently, lower obesity prevalence. Thus, the pattern of height and weight misreporting across sociodemographic groups suggests that reporting error is influenced by physiological processes (e.g.: age-related height loss), social, cultural and cognitive factors associated with body image (4,6,26,30). These errors have implications for BMI classification, because even small discrepancies in height and weight shift individuals across diagnostic thresholds (7), leading to non-trivial misclassification despite high correlations between self-reported and measured values (10–12).

Calibration of self-reported anthropometry can improve the utility of self-reported BMI in population surveillance. Models that regress measured values on self-reported data and a set of sociodemographic predictors (e.g.: age, sex, region, education) reduce systematic underestimation of BMI and align calibrated obesity prevalence with measured values (14,15,17,31). In this study, calibration almost fully corrects obesity underestimation among women and slightly overestimates obesity among men (0.8%), increasing the sensitivity of obesity classification. These findings are consistent with studies indicating that correction improves classification accuracy and the sensitivity of obesity detection relative to uncorrected BMI, although the gain is smaller for normal-weight and pre-obesity groups (14,15,17).

In this study, misreporting affects estimates of socioeconomic inequalities in obesity in a sex-specific manner, reflecting differences in BMI distributions and in reporting patterns across socioeconomic groups (11,32). Because misreporting is associated with both BMI and SEP, these relationships generate differential bias in inequality measures (12). Relative measures such as the RII are particularly sensitive to misreporting because they depend on proportional differences between groups, whereas absolute measures tend to be more stable (33).

These findings have implications for the design and interpretation of obesity surveillance and inequality monitoring. In surveys where measured anthropometry cannot be collected for all participants, incorporating small validation subsamples with both measured and self-reported data allows calibration models to reflect current misreporting patterns. Applying calibrated BMI, at least as sensitivity analysis, helps distinguish obesity socioeconomic gradients from artefacts of measurement error. As social, cultural and economic contexts and obesity prevalence evolve (34), surveillance systems should anticipate a change in misreporting patterns and periodically reassess measurement error to maintain valid and equity-relevant monitoring. Since many European surveillance systems and comparative analyses rely on self-reported BMI, similar calibration strategies could be implemented in other national health surveys with concurrent measured and self-reported anthropometry to strengthen comparability of obesity and inequality estimates across countries. In Portugal, national data continue to indicate persistent education-related obesity inequalities, particularly among women, and rising relative inequalities among men (18). Our results suggest that obesity inequalities among men may have been overestimated when based on self-reported data (33). In the absence of calibration, education-related inequalities in men may therefore be overestimated, whereas those among women may be understated, affecting the interpretability of temporal trends and cross-national comparisons.

This study has several strengths. It uses nationally representative data with concurrent measured and self-reported anthropometry collected using standardised protocols by trained fieldworkers, allowing robust estimation of misreporting patterns and the development of sex-specific calibration equations. These equations are straightforward to implement in nationally representative surveys because they rely on commonly available variables (age, region, education). The analysis accounts for the complex survey design and evaluates both absolute and relative inequality measures across multiple socioeconomic indicators, providing a comprehensive assessment of how measurement error influences obesity surveillance.

Some limitations should be considered. First, the main analyses were based on complete cases despite ∼10–11% missing values in income. If missingness relates to both SEP and obesity, complete-case analyses may under- or overestimate inequalitiesbutsensitivity analyses using multiple imputation yielded measured and calibrated BMI estimates that were broadly consistent with complete-case analyses, supporting the robustness of our findings (25,35). Second, although models adjust for key sociodemographic characteristics, residual confounding by unmeasured factors such as body image, weight-control behaviours, or cultural norms cannot be excluded (5). In additional analyses, smoking status, physical activity and marital status were included in the calibration equations, but did not improve model performance and were excluded from final models. Third, calibration equations were internally validated but developed within a single survey and time period; because because the magnitude and determinants of misreporting patterns may change across populations, survey waves and data collection modes (2), they should be revalidated before use in other settings to prevent over□or under□correction in specific subgroups. Fourth, backward variable selection yields parsimonious models with better predictive performance than more saturated specifications (14,31,36,37), but may omit variables through which SEP influences misreporting, leaving residual bias (31,36). Finally, some misclassification likely persists after calibration (17,37), particularly near BMI cut-offs, so calibrated estimates should still be interpreted cautiously and considered context-specific.

## Conclusion

Among Portuguese adults, self□reported height and weight show systematic and socially patterned misreporting that biases estimates of obesity prevalence and socioeconomic inequalities. Calibration of self□reported anthropometry using simple models incorporating age, sex, region, and education improves agreement with measured BMI, corrects the underestimation of obesity prevalence and attenuates overestimation of education-related inequalities among men. Integrating small validation subsamples and calibration procedures into national surveys would strenghten the validity and equity relevance of obesity surveillance. Where validation data are unavailable, sensitivity analyses using published calibration equations or plausible misreporting scenarios are crucial, and inequalities based on self-reported BMI should be interpreted with caution.

## Funding

This work was supported by the EEA Grants Programme, Public Health Initiatives [PT06-000088SI3]; the Fundação para a Ciência e a Tecnologia, I.P. (FCT) [grant numbers UID/4750/2025 - https://doi.org/10.54499/UID/04750/2025; LA/P/0064/2020 - https://doi.org/10.54499/UID/04750/2025];, 2023.00992.BD to B.V., CEECIND/01271/2018/CP1538/CT0002 to J.A.].

## Conflict of interest

None declared.

## Data availability

Data from the IAN-AF 2015–2016 study is publicly available and can be requested at https://www.ian-af.up.pt/en/. Sample R code used for the analyses is available from the corresponding author upon reasonable request.

## Notes

### Competing Interest Statement

The authors have declared no competing interest.

### Author Declarations

Ethics Committee/IRB of the Institute of Public Health of the University of Porto gave ethical approval for this work.

