## Supplements for "Calibrating self-reported BMI in national surveillance: impact on obesity misclassification and socioeconomic inequalities in Portugal"

### **Figure S1**. Flowchart of participant selection for the analytic sample

Total participants

(n= 6553)

Pregnant (n=61)

Age <18 (n= 2214)

Final adult analytical sample
(n= 3404)

Adult sample

(n= 4278)

Missing on:

- Self-reported weight (n= 493)
- Measured weight (n=17)
- Self-reported height (n=223)
- Measured height (n=141)

### **Section S1.** Conceptual framework: why misreporting is systematic and socially patterned

Self-reported height and weight are not only affected by random recall error and rounding/heaping but may reflect systematic, norm-oriented reporting, leading to systematic and group-specific biases. In an economic (behavioural) perspective, respondents may trade off (i) the benefit of accuracy (or the cost of misreporting) against (ii) the benefit of presenting a socially desirable body size, relative to perceived norms. A parsimonious representation is a quadratic-loss model in which individuals choose a reported value $r$(e.g., weight, height, or BMI) given the true value $w$and a reference norm $m$:

$$\max_{r}\text{ }U(r)=-a(r-w)^{2}-b(r-m)^{2},$$

where $a>0$captures the value of accuracy (or the cost of deviating from truth) and $b>0$ captures the value of social desirability (or the cost of deviating from the norm). The optimal report is a weighted average of truth and norm:

$$r=\frac{a}{a+b}w+\frac{b}{a+b}m,$$

implying that reporting error $e=r-w$ systematically pulls reports toward the norm $m$, with the strength of this “pull” increasing in $b/(a+b)$. We use this as a conceptual device rather than a structural model to be estimated: importantly, the reference value m and/or the relative weight placed on accuracy and desirability $\frac{a}{(a+b)}$ and $\frac{b}{(a+b)}$ may vary across contexts and socioeconomic group.

This conceptualization generates testable implications that help interpret why misreporting varies with measured BMI, age, and socioeconomic position (SEP). Importantly, if misreporting is correlated with SEP, then the resulting measurement error is differential rather than classical and can bias both prevalence estimates and inequality measures constructed from self-reported BMI.

Hypotheses

H1 (Systematic direction): Mean reporting errors are directionally patterned (e.g., weight underreported and height overreported) because norms typically favor lower weight and (to a lesser degree) greater height.

H2 (BMI gradient/distance-to-norm): The magnitude of reporting error increases with measured BMI (and may be nonlinear), consistent with a “pull” toward socially desirable reference values.

H3 (Heterogeneity by SEP): The reporting process is socially patterned: the reference value m and/or the relative weight placed on desirability versus accuracy (b/(a+b) versus a/(a+b)) varies across SEP groups (e.g., education or income). Consequently, reporting error differs systematically by SEP, potentially in both direction and magnitude, reflecting differences in perceived norms and stigma, social comparison, health awareness, and response styles.

H4 (Age, awareness, and physiological change): Misreporting increases with age through two distinct channels. First, awareness of current anthropometrics declines: older adults are less likely to have recently measured their weight and may recall outdated values. Second, height is subject to age-related shrinkage; if respondents report their historical maximum height, self-reported height increasingly exceeds measured height with age. Additional rounding/heaping with age may widen discrepancies. As a result, absolute error rises with age, and age-related misreporting is expected to be strongest among individuals with higher measured BMI, consistent with stronger norm pressure and greater uncertainty about current weight.

H5 (Implications for inequalities): Because misreporting is correlated with both BMI and SEP, inequality estimates based on self-reported BMI are biased relative to those based on measured BMI; calibration reduces but may not eliminate this bias, and the direction of change depends on how misreporting differs across SEP groups within sex. Calibration is therefore not a neutral rescaling: it can alter between-group contrasts and thus PD/PR/SII/RII.

### **Table S1**. Comparison between complete and incomplete cases among included participants (N=3404)

|  |  | **Complete (n, %)** 3,072 (90.2) | **Incomplete (n, %)** 332 (9.8) | ***p*** |
| --- | --- | --- | --- | --- |
| **Age, Mean (SD)** |  | 46.6 (15.9) | 43.2 (19.9) | <0.001 |
| **Sex** |  |  |  | 0.150 |
| Women |  | 1,546 (50.3) | 181 (54.5) |  |
| Men |  | 1,526 (49.7) | 151 (45.5) |  |
| **Region of residence** |  |  |  | 0.200 |
| North |  | 558 (18.2) | 65 (19.6) |  |
| Center |  | 486 (15.8) | 56 (16.9) |  |
| Lisbon Metropolitan Area |  | 388 (12.6) | 49 (14.8) |  |
| Alentejo |  | 361 (11.8) | 32 (9.6) |  |
| Algarve |  | 430 (14.0) | 30 (9.0) |  |
| Autonomous Region of Madeira |  | 416 (13.5) | 50 (15.1) |  |
| Autonomous Region of Azores |  | 433 (14.1) | 50 (15.1) |  |
| **Education level** |  |  |  | <0.001 |
| Low |  | 1487 (49.1) | 177 (54.4) |  |
| Intermediate |  | 797 (25.9) | 100 (30.8) |  |
| High |  | 768 (25.0) | 48 (14.8) |  |
| *Missing* |  | 0 | 7 (2.1) |  |
| **Income quintile** |  | 3.4 (1.9) | 3.1 (2.8) | 0.200 |
| *Missing* |  | 0 | 323 (10.8) |  |
| **Employment status** |  |  |  | <0.001 |
| Employed |  | 1,839 (59.9) | 130 (40.1) |  |
| Other employment situation |  | 887 (28.9) | 149 (46.0) |  |
| Unemployed |  | 346 (11.3) | 45 (13.9) |  |
| *Missing* |  | 0 | 8 (0.2) |  |
| Abbreviations: SD, standard deviation. P values obtained from Wilcoxon rank-sum test and Pearson’s chi-squared test. | | | | |

### **Table S2**. Unweighted participants’ characteristics by sex at birth

|  | **Women** **(n, %)** N = 1,727^1^ | **Men (n, %)**  N = 1,677^1^ |
| --- | --- | --- |
| **Age. Mean (SD)** | 45.1 (15.9) | 47.6 (16.8) |
| **Region of residence** |  |  |
| North | 308 (17.8) | 315 (18.8) |
| Center | 263 (15.2) | 279 (16.6) |
| Lisbon Metropolitan Area | 239 (13.8) | 198 (11.8) |
| Alentejo | 201 (11.6) | 192 (11.4) |
| Algarve | 240 (13.9) | 220 (13.1) |
| Autonomous Region of Madeira | 242 (14.0) | 224 (13.4) |
| Autonomous Region of Azores | 234 (13.5) | 249 (14.8) |
| **Marital status** |  |  |
| Single | 464 (26.9) | 445 (26.5) |
| Divorced | 147 (8.5) | 119 (7.1) |
| Widowed | 110 (6.4) | 44 (2.6) |
| Married or in a civil union | 1,004 (58.2) | 1,069 (63.7) |
| **Education** |  |  |
| Low | 773 (44.9) | 911 (54.4) |
| Intermediate | 469 (27.2) | 428 (25.6) |
| High | 481 (27.9) | 335 (20.0) |
| **Employment status** |  |  |
| Employed | 1,006 (58.5) | 963 (57.4) |
| Other employment situation | 507 (29.5) | 529 (31.5) |
| Unemployed | 206 (12.0) | 185 (11.0) |
| **Income quintile** |  |  |
| Low | 628 (40.4%) | 537 (35.1) |
| Intermediate | 647 (41.7%) | 622 (40.7) |
| High | 278 (17.9%) | 369 (24.1) |
| **Weight (kg), mean (SD)** |  |  |
| Self-reported | 67.2 (13.4) | 79.4 (13.5) |
| Measured | 67.9 (13.8) | 79.9 (13.8) |
| **Height (cm), mean (SD)** |  |  |
| Self-reported | 160.5 (6.7) | 172.7 (7.3) |
| Measured | 158.6 (6.7) | 171.5 (7.4) |
| **BMI (kg/m^2^), mean (SD)** |  |  |
| Self-reported | 26.1 (5.2) | 26.6 (4.2) |
| Measured | 27.1 (5.6) | 27.2 (4.4) |

Abbreviations: N, number of participants; n, number in subgroup; SD, standard deviation; BMI, body mass index; kg, kilograms; cm, centimetres.

### **Figure S2**. Bland–Altman plots comparing self-reported and measured **weight and height** by sex at birth

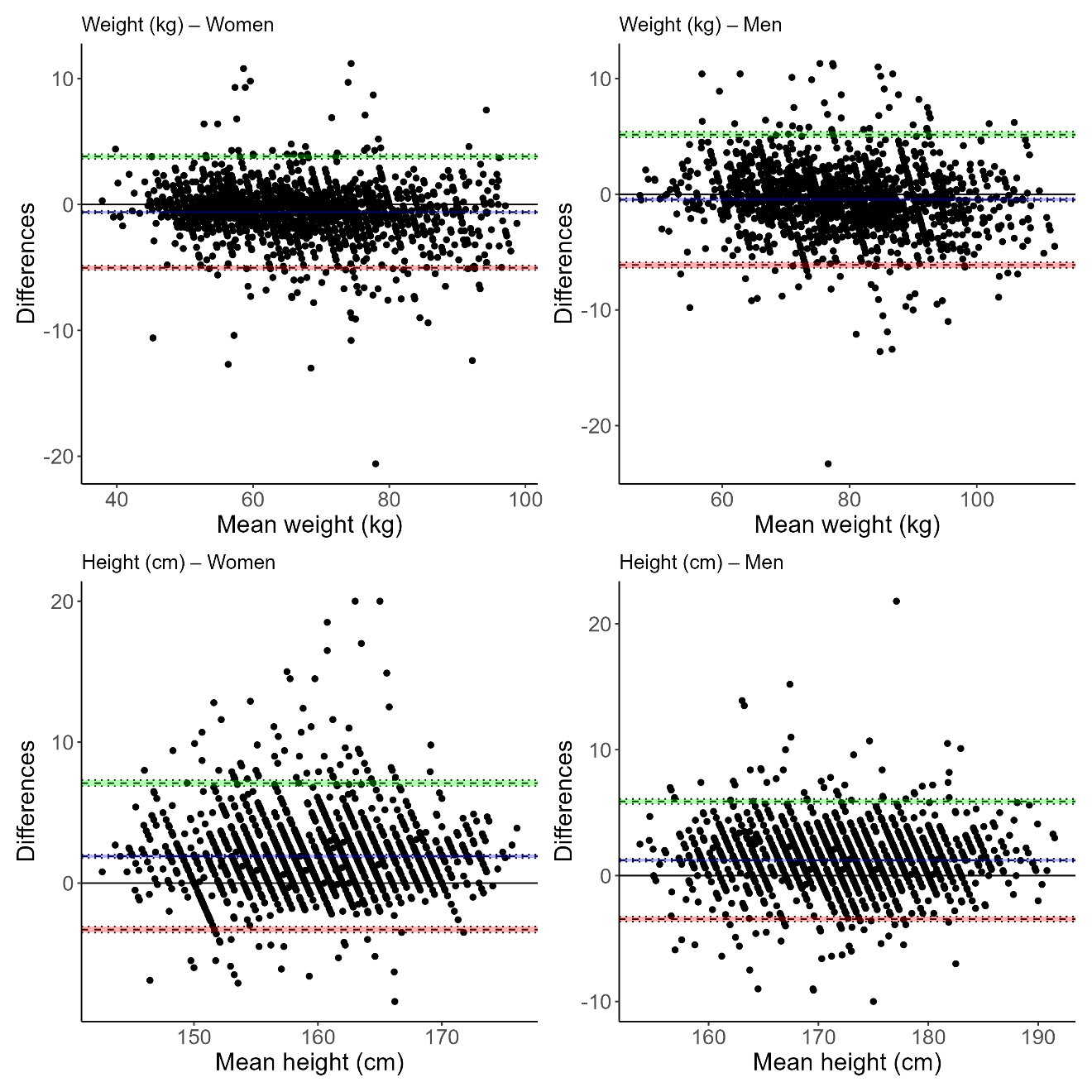

Note: Difference = self-reported (height or weight) – measured (height or weight).

Difference > 0, overreport; Difference < 0, underreport.

### **Figure S3**. Age-related bias in self-reported **height, weight and BMI** by sex and BMI category

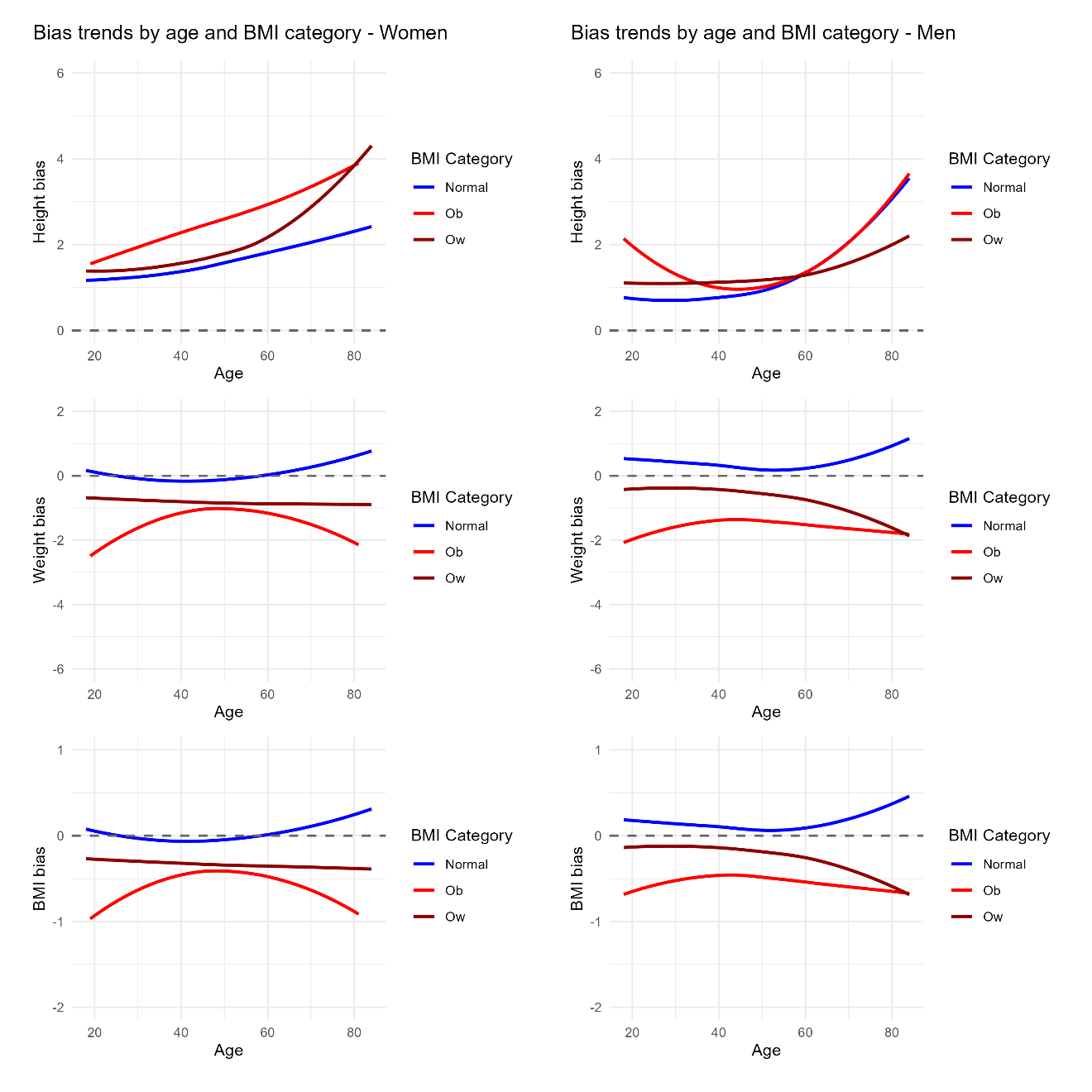

Abbreviations: BMI, body mass index; Ob, obesity; Ow, Pre-obesity; Age is described in years.

### **Figure S4.** Age‑related misreporting of height, weight, and BMI by sex

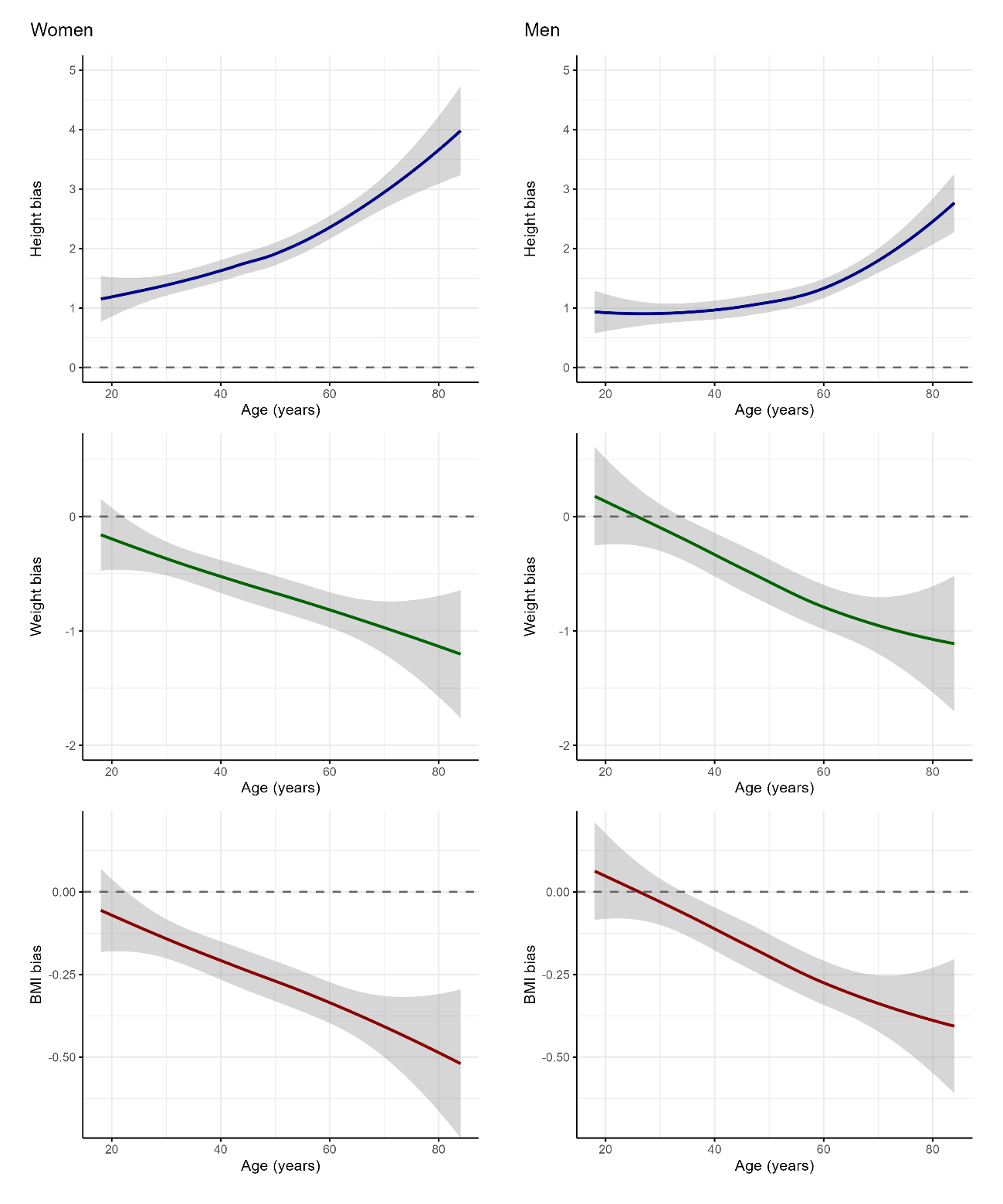

### **Table S3**. Determinants of height (± 1 cm) and weight (± 2 kg) bias (no bias as reference) relative to measured values using adjusted sex-stratified survey-weighted multinomial regression model

|  | **Height** | | | | **Weight** | | | |
| --- | --- | --- | --- | --- | --- | --- | --- | --- |
|  | *Women* | | *Men* | | *Women* | | *Men* | |
|  | Underreport vs  no bias | Overreport vs  no bias | Underreport vs  no bias | Overreport vs  no bias | Underreport vs  no bias | Overreport vs  no bias | Underreport vs  no bias | Overreport vs  no bias |
|  | OR (95%CI) | | | | | | | |
| Age (years) | 1.06 (1.03–1.10) | 1.03 (1.02–1.04) | 1.00 (0.98–1.02) | 1.00 (0.99–1.02) | 0.99 (0.97–1.01) | 1.00 (0.98–1.02) | 1.01 (0.99–1.03) | 0.98 (0.96–1.00) |
| **Region of residence** |  |  |  |  |  |  |  |  |
| North | 1.08 (0.59–1.99) | 1.84 (1.42–2.39) | 1.8 (1.10–2.96) | 1.29 (0.96–1.72) | 0.99 (0.72–1.36) | 0.85 (0.52–1.38) | 1.12 (0.80–1.56) | 0.73 (0.44–1.24) |
| Center | 0.72 (0.41–1.24) | 1.00 (0.71–1.39) | 1.39 (0.92–2.10) | 1.32 (0.93–1.88) | 1.21 (0.86–1.71) | 1.22 (0.68–2.19) | 0.92 (0.68–1.25) | 1.35 (0.93–1.97) |
| Lisbon Metropolitan Area | 0.84 (0.39–1.84) | 0.94 (0.67–1.31) | 0.59 (0.31–1.11) | 1.08 (0.73–1.58) | 0.98 (0.62–1.55) | 0.54 (0.29–1.03) | 0.78 (0.53–1.16) | 0.91 (0.57–1.45) |
| Alentejo | 0.50 (0.16–1.50) | 1.00 (0.62–1.62) | 0.82 (0.45–1.49) | 1.24 (0.84–1.82) | 0.69 (0.49–0.96) | 1.11 (0.62–1.98) | 0.64 (0.37–1.10) | 1.67 (1.23–2.28) |
| Algarve | 0.76 (0.41–1.41) | 0.59 (0.46–0.74) | 0.70 (0.48–1.02) | 0.70 (0.56–0.88) | 0.76 (0.50–1.16) | 1.23 (0.73–2.05) | 1.46 (1.14–1.86) | 0.59 (0.39–0.89) |
| Autonomous Region of Madeira | 2.05 (1.20–3.51) | 1.02 (0.71–1.45) | 1.60 (1.17–2.20) | 0.93 (0.66–1.32) | 1.12 (0.82–1.54) | 0.97 (0.48–1.96) | 1.25 (0.89–1.77) | 1.03 (0.75–1.41) |
| Autonomous Region of Azores | 0.73 (0.49-1.10) | 0.68 (0.51-0.90) | 0.73 (0.49–1.10) | 0.68 (0.51–0.90) | 1.45 (0.97–2.18) | 1.35 (0.58–3.14) | 1.06 (0.79–1.42) | 1.09 (0.79–1.51) |
| **Education level** |  |  |  |  |  |  |  |  |
| Low | 0.70 (0.24–2.07) | 0.89 (0.53–1.48) | 1.36 (0.5–3.68) | 1.1 (0.62–1.97) | 0.84 (0.48–1.47) | 0.74 (0.41–1.36) | 0.98 (0.54–1.78) | 1.86 (0.96–3.58) |
| Intermediate | 1.00 (0.45–2.2) | 1.20 (0.75–1.91) | 1.2 (0.55–2.62) | 1.1 (0.68–1.79) | 1.11 (0.62–1.99) | 0.90 (0.46–1.75) | 1.03 (0.51–2.08) | 1.70 (0.88–3.30) |
| High | Ref. | Ref. | Ref. | Ref. | Ref. | Ref. | Ref. | Ref. |
| **Income level** |  |  |  |  |  |  |  |  |
| Low | 2.06 (0.62–6.90) | 0.94 (0.59–1.50) | 2.83 (1.35–5.92) | 1.13 (0.69–1.86) | 1.27 (0.73–2.21) | 1.64 (0.74–3.62) | 1.03 (0.54–1.95) | 2.53 (1.24–5.15) |
| Intermediate | 2.72 (1.09–6.79) | 1.03 (0.67–1.60) | 1.47 (0.85–2.55) | 0.94 (0.62–1.44) | 0.68 (0.41–1.14) | 1.22 (0.56–2.67) | 0.77 (0.45–1.32) | 1.53 (0.86–2.71) |
| High | Ref. | Ref. | Ref. | Ref. | Ref. | Ref. | Ref. | Ref. |
| **Employment status** |  |  |  |  |  |  |  |  |
| Employed | Ref. | Ref. | Ref. | Ref. | Ref. | Ref. | Ref. | Ref. |
| Other employment situation | 0.62 (0.22–1.79) | 0.78 (0.52–1.17) | 0.61 (0.29–1.29) | 0.96 (0.63–1.45) | 1.70 (1.03–2.79) | 1.37 (0.81–2.32) | 1.19 (0.67–2.13) | 0.55 (0.30–1.01) |
| Unemployed | 1.84 (0.74–4.58) | 0.88 (0.54–1.44) | 0.52 (0.23–1.20) | 0.62 (0.33–1.16) | 1.70 (0.92–3.13) | 1.94 (0.73–5.16) | 2.06 (1.06–3.98) | 1.28 (0.61–2.72) |
| **BMI category** |  |  |  |  |  |  |  |  |
| Normal weight | Ref. | Ref. | Ref. | Ref. | Ref. | Ref. | Ref. | Ref. |
| Pre-obesity | 1.33 (0.64–2.77) | 0.98 (0.66–1.47) | 0.92 (0.49–1.73) | 1.57 (1.04–2.38) | 2.37 (1.42–3.95) | 0.38 (0.20–0.74) | 3.51 (2.07–5.94) | 0.65 (0.36–1.18) |
| Obesity | 0.39 (0.14–1.1) | 1.64 (0.98–2.74) | 0.95 (0.46–1.98) | 1.14 (0.73–1.80) | 5.06 (2.90–8.84) | 1.03 (0.44–2.39) | 7.47 (4.18–13.33) | 0.47 (0.22–0.97) |

Abbreviations: OR, odds ratio; CI, confidence interval; BMI, body mass index; Ref, reference group.

### **Table S4**. Comparison of measured and self-reported **height** by sex and sociodemographic characteristics

|  | | *Women* | | | | | *Men* | | | | |
| --- | --- | --- | --- | --- | --- | --- | --- | --- | --- | --- | --- |
|  |  | Measured | Self-reported | Difference | p-value | | Measured | Self-reported | Difference | p-value | |
|  |  | Mean (SD) | | | within | between | Mean (SD) | | | within | between |
| **Age group (years)** | 18-19 | 162.2 (1.5) | 163.2 (1.5) | 1.04 (0.24) | <0.001 | <0.001 | 175.6 (1.1) | 175.9 (1.1) | 0.23 (0.41) | <0.001 | <0.001 |
|  | 20-29 | 161.5 (0.6) | 162.8 (0.6) | 1.27 (0.16) | <0.001 |  | 176.1 (0.5) | 177.0 (0.6) | 0.86 (0.18) | <0.001 |  |
|  | 30-39 | 160.7 (0.4) | 162.6 (0.4) | 1.89 (0.19) | <0.001 |  | 174.8 (0.4) | 176.0 (0.4) | 1.20 (0.19) | <0.001 |  |
|  | 40-49 | 158.5 (0.4) | 160.4 (0.4) | 1.88 (0.18) | <0.001 |  | 173.0 (0.6) | 174.0 (0.6) | 0.93 (0.19) | <0.001 |  |
|  | 50-59 | 157.0 (0.4) | 159.1 (0.4) | 2.07 (0.23) | <0.001 |  | 169.4 (0.6) | 170.4 (0.6) | 1.00 (0.15) | <0.001 |  |
|  | 60-69 | 155.3 (0.4) | 158.1 (0.4) | 2.73 (0.2) | <0.001 |  | 167.7 (0.6) | 169.1 (0.6) | 1.42 (0.31) | <0.001 |  |
|  | 70-79 | 152.9 (0.7) | 156.6 (0.9) | 3.68 (0.54) | <0.001 |  | 167.1 (0.7) | 168.8 (0.7) | 1.71 (0.31) | <0.001 |  |
|  | ≥80 | 152.4 (1.2) | 155.5 (2.1) | 3.09 (1.64) | 0.102 |  | 164 (1.3) | 166.5 (1.2) | 2.55 (0.43) | <0.001 |  |
| **Region of residence** | North | 157.4 (0.5) | 159.8 (0.5) | 2.45 (0.22) | <0.001 | 0.001 | 171.6 (0.4) | 172.7 (0.5) | 1.07 (0.21) | <0.001 | 0.210 |
|  | Center | 158.2 (0.4) | 160.0 (0.4) | 1.81 (0.19) | <0.001 |  | 171.3 (0.7) | 172.4 (0.6) | 1.11 (0.17) | <0.001 |  |
|  | Lisbon Metropolitan Area | 159.1 (0.5) | 160.9 (0.6) | 1.86 (0.15) | <0.001 |  | 172.0 (0.6) | 173.3 (0.6) | 1.23 (0.17) | <0.001 |  |
|  | Alentejo | 158.5 (0.4) | 160.8 (0.3) | 2.30 (0.29) | <0.001 |  | 171.0 (0.6) | 172.4 (0.6) | 1.44 (0.19) | <0.001 |  |
|  | Algarve | 158.8 (0.5) | 160.1 (0.4) | 1.32 (0.16) | <0.001 |  | 172.1 (0.4) | 173 (0.5) | 0.86 (0.11) | <0.001 |  |
|  | Autonomous Region of Madeira | 158.7 (0.3) | 160.2 (0.3) | 1.50 (0.23) | 0.001 |  | 171.1 (0.9) | 172.0 (1) | 0.96 (0.27) | 0.015 |  |
|  | Autonomous Region of Azores | 158.6 (0.6) | 160.8 (0.4) | 2.17 (0.18) | <0.001 |  | 172.1 (0.5) | 173.2 (0.5) | 1.11 (0.17) | 0.001 |  |
| **BMI category** | Normal weight | 159.8 (0.3) | 161.4 (0.3) | 1.61 (0.14) | <0.001 | <0.001 | 172.9 (0.4) | 173.7 (0.4) | 0.87 (0.15) | <0.001 | 0.112 |
|  | Pre-obesity | 158.1 (0.4) | 160 (0.4) | 1.88 (0.18) | <0.001 |  | 171.4 (0.4) | 172.7 (0.4) | 1.27 (0.14) | <0.001 |  |
|  | Obesity | 155.6 (0.4) | 158.7 (0.5) | 3.06 (0.22) | <0.001 |  | 170.1 (0.5) | 171.3 (0.6) | 1.24 (0.22) | <0.001 |  |
| **Education level** | Low | 156.1 (0.3) | 158.6 (0.3) | 2.51 (0.19) | <0.001 | 0.001 | 169.4 (0.3) | 170.6 (0.4) | 1.11 (0.16) | <0.001 | 0.870 |
|  | Intermediate | 159.5 (0.5) | 161.3 (0.5) | 1.81 (0.16) | <0.001 |  | 173.2 (0.5) | 174.4 (0.5) | 1.20 (0.16) | <0.001 |  |
|  | High | 160.3 (0.4) | 161.9 (0.4) | 1.62 (0.11) | <0.001 |  | 174.7 (0.4) | 175.8 (0.4) | 1.09 (0.14) | <0.001 |  |
| **Income level** | Low | 156.9 (0.4) | 159.3 (0.3) | 2.42 (0.2) | <0.001 | 0.039 | 169.5 (0.4) | 170.5 (0.4) | 1.00 (0.21) | <0.001 | 0.572 |
|  | Intermediate | 158.6 (0.3) | 160.4 (0.3) | 1.82 (0.1) | <0.001 |  | 172 (0.4) | 173.2 (0.4) | 1.13 (0.14) | <0.001 |  |
|  | High | 159.5 (0.4) | 161.3 (0.5) | 1.87 (0.17) | <0.001 |  | 173.2 (0.5) | 174.5 (0.5) | 1.24 (0.1) | <0.001 |  |
| **Employment status** | Employed | 159.3 (0.2) | 161.2 (0.3) | 1.92 (0.1) | <0.001 | <0.001 | 172.9 (0.4) | 174 (0.4) | 1.07 (0.11) | <0.001 | 0.086 |
|  | Other employment situation | 156.4 (0.4) | 159.1 (0.4) | 2.66 (0.23) | <0.001 |  | 169 (0.4) | 170.3 (0.5) | 1.39 (0.16) | <0.001 |  |
|  | Unemployed | 157.7 (0.7) | 159.1 (0.7) | 1.36 (0.24) | <0.001 |  | 171.8 (0.8) | 172.6 (0.8) | 0.84 (0.24) | <0.001 |  |

Abbreviations: BMI, body mass index; SD, standard deviation.

### **Table S5**. Comparison of measured and self-reported **weight** by sex and sociodemographic characteristics

|  |  | **Women** | | | | | **Men** | | | | |
| --- | --- | --- | --- | --- | --- | --- | --- | --- | --- | --- | --- |
|  |  | *Measured* | *Self-reported* | *Difference* | *p-value* | | *Measured* | *Self-reported* | *Difference* | *p-value* | |
|  |  | Mean (SD) | | | within | between | Mean (SD) | | | within | between |
| **Age group (years)** | 18-19 | 61.5 (2.1) | 61.5 (2.2) | -0.03 (0.23) | 0.887 | 0.092 | 72.7 (1.8) | 72.3 (1.6) | -0.37 (0.54) | 0.498 | <0.001 |
|  | 20-29 | 60.9 (0.9) | 60.3 (0.9) | -0.61 (0.17) | 0.001 |  | 73.9 (1.1) | 74.4 (1.0) | 0.52 (0.3) | 0.087 |  |
|  | 30-39 | 65.3 (0.8) | 64.9 (0.8) | -0.45 (0.2) | 0.026 |  | 80.5 (1) | 80.5 (0.9) | -0.04 (0.21) | 0.862 |  |
|  | 40-49 | 66.6 (0.9) | 65.9 (0.9) | -0.72 (0.17) | 0.000 |  | 81.5 (1.3) | 81.2 (1.3) | -0.31 (0.28) | 0.259 |  |
|  | 50-59 | 68.3 (0.8) | 67.5 (0.7) | -0.77 (0.17) | 0.000 |  | 79.3 (1.2) | 78.5 (1.1) | -0.8 (0.23) | 0.001 |  |
|  | 60-69 | 70.6 (1.2) | 69.9 (1.2) | -0.69 (0.22) | 0.002 |  | 78.3 (1.4) | 77.3 (1.3) | -1.09 (0.21) | 0.000 |  |
|  | 70-79 | 70.6 (1.6) | 70.0 (1.3) | -0.54 (0.73) | 0.464 |  | 81.1 (1.9) | 79.5 (2.0) | -1.63 (0.62) | 0.011 |  |
|  | ≥80 | 63.3 (4.3) | 61.9 (4.1) | -1.42 (0.5) | 0.021 |  | 74.2 (2.3) | 72.8 (2.2) | -1.37 (0.33) | 0.000 |  |
| **Region** | North | 65.9 (0.7) | 65.4 (0.7) | -0.53 (0.17) | 0.005 | 0.004 | 79.5 (0.7) | 78.6 (0.7) | -0.82 (0.15) | 0.000 | 0.017 |
|  | Center | 66.3 (1.1) | 65.6 (1.1) | -0.77 (0.17) | 0.000 |  | 77.7 (1.4) | 77.6 (1.4) | -0.08 (0.19) | 0.701 |  |
|  | Lisbon Metropolitan Area | 67.3 (0.7) | 66.5 (0.7) | -0.8 (0.17) | 0.000 |  | 77.9 (1.4) | 77.6 (1.4) | -0.25 (0.2) | 0.227 |  |
|  | Alentejo | 66.8 (0.9) | 66.6 (0.8) | -0.15 (0.1) | 0.169 |  | 81 (1.4) | 80.8 (1.5) | -0.2 (0.47) | 0.677 |  |
|  | Algarve | 67.2 (0.5) | 66.6 (0.4) | -0.58 (0.14) | 0.001 |  | 78.3 (0.4) | 77.4 (0.5) | -0.85 (0.19) | 0.001 |  |
|  | Autonomous Region of Madeira | 67.4 (0.9) | 66.6 (0.9) | -0.83 (0.18) | 0.006 |  | 77.8 (1.8) | 77.4 (1.7) | -0.42 (0.17) | 0.062 |  |
|  | Autonomous Region of Azores | 68.6 (0.7) | 68.0 (0.8) | -0.54 (0.13) | 0.009 |  | 80.4 (0.8) | 79.5 (0.9) | -0.82 (0.19) | 0.008 |  |
| **BMI category** | Normal weight | 56.8 (0.3) | 56.7 (0.3) | -0.05 (0.12) | 0.697 | <0.001 | 67.6 (0.5) | 68.2 (0.5) | 0.64 (0.17) | 0.000 | <0.001 |
|  | Overweight | 68.8 (0.4) | 67.9 (0.4) | -0.89 (0.11) | 0.000 |  | 80.6 (0.5) | 79.9 (0.5) | -0.74 (0.16) | 0.000 |  |
|  | Obesity | 80.6 (0.5) | 79.3 (0.5) | -1.31 (0.18) | 0.000 |  | 93.6 (0.7) | 91.8 (0.8) | -1.83 (0.26) | 0.000 |  |
| **Education level** | Low | 69.7 (0.6) | 68.9 (0.5) | -0.78 (0.13) | 0.000 | 0.160 | 78.5 (0.7) | 78.1 (0.6) | -0.43 (0.18) | 0.019 | 0.066 |
|  | Intermediate | 64.9 (0.7) | 64.3 (0.7) | -0.58 (0.16) | 0.000 |  | 77.8 (1) | 77.6 (0.9) | -0.23 (0.21) | 0.269 |  |
|  | High | 63.1 (0.7) | 62.6 (0.7) | -0.49 (0.1) | 0.000 |  | 80.3 (1) | 79.4 (1) | -0.88 (0.17) | 0.000 |  |
| **Income level** | Low | 68.3 (0.6) | 67.4 (0.6) | -0.9 (0.13) | <0.001 | 0.137 | 78 (0.8) | 77.5 (0.8) | -0.44 (0.27) | 0.109 | 0.006 |
|  | Intermediate | 66.7 (0.6) | 66.1 (0.6) | -0.56 (0.11) | <0.001 |  | 78.3 (0.8) | 78 (0.7) | -0.31 (0.13) | 0.018 |  |
|  | High | 63.6 (0.8) | 63 (0.8) | -0.58 (0.14) | <0.001 |  | 81.8 (1) | 80.8 (1) | -1.05 (0.17) | 0.000 |  |
| **Employment status** | Employed | 66.1 (0.6) | 65.5 (0.6) | -0.61 (0.08) | 0.000 | 0.665 | 80.3 (0.6) | 80 (0.6) | -0.38 (0.11) | 0.001 | 0.028 |
|  | Unemployed | 65.3 (1) | 64.5 (1) | -0.88 (0.3) | 0.004 |  | 74.4 (1.7) | 74.5 (1.6) | 0.07 (0.44) | 0.872 |  |
|  | Other employment situation | 68 (0.7) | 67.4 (0.7) | -0.61 (0.2) | 0.003 |  | 77.4 (0.9) | 76.5 (0.8) | -0.91 (0.19) | 0.000 |  |

Abbreviations: BMI, body mass index; SD, standard deviation.

### **Table S6**. Calibrated height and weight coefficients by sex at birth

|  |  | **Height** | | **Weight** | |
| --- | --- | --- | --- | --- | --- |
|  |  | Women | Men | Women | Men |
|  |  | β (95% CI) | β (95% CI) | β (95% CI) | β (95% CI) |
| **Intercept** |  | 24.678 (19.689–29.667) | 16.342 (11.091–21.593) | -0.260 (-1.222; 0.703) | -1.597 (-3.242–0.048) |
| **Self-reported Height/Weight** | | 0.848 (0.819–0.878) | 0.91 (0.882–0.938) | 1.009 (0.992, 1.023) | 1.013 (0.992–1.034) |
| **Age (years)** | | -0.053 (-0.065–-0.04) | -0.035 (-0.048–-0.021) | 0.003 (-0.013, 0.019) | 0.043 (0.03–0.057) |
| **Region of residence** | North | Ref | Ref | Ref | Ref |
|  | Center | 0.728 (0.154–1.303) | -0.039 (-0.576–0.498) | 0.237 (-0.25–0.725) | -0.778 (-1.281–-0.275) |
|  | Lisbon M.A. | 0.685 (0.188–1.183) | -0.161 (-0.66–0.339) | 0.289 (-0.202–0.78) | -0.665 (-1.225–-0.106) |
|  | Alentejo | 0.43 (-0.245–1.105) | -0.378 (-0.91–0.155) | -0.409 (-0.814–-0.004) | -0.725 (-1.657–0.208) |
|  | Algarve | 1.134 (0.554–1.715) | 0.148 (-0.303–0.598) | 0.052 (-0.385–0.49) | 0.141 (-0.333–0.616) |
|  | Autonomous Region of Madeira | 0.956 (0.298–1.613) | -0.026 (-0.649–0.597) | 0.297 (-0.203–0.797) | -0.328 (-0.778–0.121) |
|  | Autonomous Region of Azores | 0.470 (-0.205–1.144) | -0.07 (-0.625–0.485) | 0.006 (-0.451–0.463) | 0.101 (-0.411–0.614) |
| **Education level** | Low | -0.656 (-1.018–-0.295) | -0.153 (-0.603–0.297) | 0.223 (-0.093–0.538) | -0.949 (-1.464–-0.434) |
|  | Intermediate | -0.445 (-0.768–-0.122) | -0.36 (-0.805–0.086) | 0.082 (-0.306–0.469) | -0.512 (-1.067–0.043) |
|  | High | Ref. | Ref. | Ref. | Ref. |

Example:

To facilitate implementation, we provide worked examples of the computation in Supplementary Material Box 1 (e.g. for one male and one female participant with different regions and education levels). Users should ensure that the coding of region and education in their data matches the reference categories and indicator coding used in our models; otherwise, the calibration coefficients will not be directly applicable.

### **Table S7**. Survey-weighted mean and standard error of measured, self-reported and calibrated anthropometric variables by sex

|  | **Weight (kg)** | | | **Height (cm)** | | | **BMI (kg/m²)** | | |
| --- | --- | --- | --- | --- | --- | --- | --- | --- | --- |
|  | *Mean (SE)* | | | | | | | | |
|  | Measured | Self-reported | Calibrated | Measured | Self-reported | Calibrated | Measured | Self-reported | Calibrated |
| **Women** | 66.49 (0.41) | 65.88 (0.41) | 66.52 (0.41) | 158.13 (0.26) | 160.20 (0.26) | 158.14 (0.24) | 26.68 (0.18) | 26.43 (0.18) | 26.67 (0.18) |
| **Men** | 78.61 (0.53) | 78.17 (0.53) | 78.64 (0.53) | 171.60 (0.28) | 172.72 (0.28) | 171.59 (0.27) | 26.71 (0.17) | 26.56 (0.17) | 26.73 (0.17) |

Abbreviations: BMI, body mass index; SE, standard error

### **Tabe S8**. Weighted agreement between measured and calibrated BMI categories by sex at birth

|  | | | **Calibrated BMI (%)** | | |
| --- | --- | --- | --- | --- | --- |
|  |  |  | *Normal weight* | *Pre-obesity* | *Obesity* |
| **Measured BMI (%)** | **Women** | *Normal weight* | 90.5 | 8.5 | 1.0 |
|  |  | *Pre-obesity* | 5.0 | 86.2 | 8.8 |
|  |  | *Obesity* | 0.0 | 13.2 | 86.8 |
|  | **Men** | *Normal weight* | 89.0 | 10.0 | 0.9 |
|  |  | *Pre-obesity* | 10.8 | 81.0 | 8.2 |
|  |  | *Obesity* | 0.6 | 13.9 | 85.5 |

### **Table S9**. Weighted approximation of Lin’s concordance correlation coefficient for measured, self-reported and calibrated anthropometric data

|  | | **Weight** | | **Height** | | **BMI** | |
| --- | --- | --- | --- | --- | --- | --- | --- |
|  |  | Calibrated | Self-reported | Calibrated | Self-reported | Calibrated | Self-reported |
| Women | CCC | **0.981** | 0.979 | **0.922** | 0.867 | **0.981** | 0.979 |
|  | Mean difference* | **0.021** | -0.630 | **-0.012** | 2.060 | **0** | -0.255 |
| Men | CCC | **0.974** | 0.970 | **0.949** | 0.935 | **0.970** | 0.942 |
|  | Mean difference* | **0.030** | -0.483 | **-0.003** | 0.130 | **0.009** | -0.498 |

Abbreviations: *According to measured values. BMI: Body mass index; CCC: Concordance Correlation Coefficient

### **Table S10**. Socioeconomic inequalities in obesity by BMI assessment method under multiple imputations (MICE)

|  |  | **Education** | | **Income** | | **Occupation** | |
| --- | --- | --- | --- | --- | --- | --- | --- |
|  | **BMI definition** | **PD (95%CI)** | **PR (95%CI)** | **PD (95%CI)** | **PR (95%CI)** | **PD (95%CI)** | **PR (95%CI)** |
| **Women** | *Measured* | 26.0 (19.1, 32.9) | 3.21 (2.01, 5.15) | 18.7 (10.2, 27.1) | 2.40 (1.41, 4.09) | 1.2 (-9.2, 11.7) | 1.06 (0.64, 1.78) |
|  | *Self-reported* | 19.8 (13.3, 26.3) | 3.35 (2.01, 5.60) | 14.4 (7.0, 21.9) | 2.45 (1.37, 4.38) | -1.1 (-9.1, 6.8) | 0.93 (0.53, 1.62) |
|  | *Calibrated* | 28.6 (22.2, 34.9) | 4.07 (2.62, 6.34) | 18.7 (10.3, 27.2) | 2.58 (1.53, 4.37) | 2.5 (-6.4, 11.3) | 1.13 (0.74, 1.75) |
| **Men** | *Measured* | 8.2 (1.9, 14.6) | 1.48 (1.07, 2.07) | 5.0 (-3.1, 13.1) | 1.23 (0.86, 1.76) | -3.0 (-11.7, 5.8) | 0.85 (0.50, 1.43) |
|  | *Self-reported* | 9.2 (4.5, 13.9) | 1.82 (1.28, 2.57) | 7.6 (0.7, 14.6) | 1.56 (1.02, 2.38) | -3.8 (-12.7, 5.0) | 0.77 (0.40, 1.48) |
|  | *Calibrated* | 7.6 (1.4, 13.8) | 1.44 (1.04, 2.00) | 8.9 (0.8, 17.0) | 1.47 (1.02, 2.10) | -7.1 (-16.1, 1.9) | 0.65 (0.34, 1.24) |
|  |  | **SII** | **RII** | **SII** | **RII** | **SII** | **RII** |
| **Women** | *Measured* | 42.0 (31.3, 52.8) | 6.64 (3.54, 12.46) | 25.6 (14.0, 37.2) | 2.92 (1.71, 5.01) | 25.1 (12.3, 38.0) | 2.72 (1.68, 4.40) |
|  | *Self-reported* | 32.5 (22.0, 42.9) | 7.90 (3.75, 16.64) | 20.9 (10.4, 31.5) | 3.43 (1.76, 6.68) | 11.4 (-0.6, 23.3) | 1.88 (1.00, 3.54) |
|  | *Calibrated* | 46.2 (36.4, 56.1) | 9.75 (5.41, 17.59) | 25.5 (13.0, 38.0) | 3.11 (1.73, 5.60) | 23.6 (10.4, 36.7) | 2.69 (1.58, 4.59) |
| **Men** | *Measured* | 15.7 (5.2, 26.1) | 2.21 (1.26, 3.88) | 8.5 (-2.7, 19.7) | 1.51 (0.86, 2.65) | 6.5 (-4.9, 17.9) | 1.36 (0.80, 2.31) |
|  | *Self-reported* | 16.4 (8.0, 24.7) | 2.98 (1.62, 5.48) | 11.5 (1.7, 21.3) | 2.06 (1.09, 3.91) | -4.1 (-17.0, 8.8) | 0.77 (0.34, 1.77) |
|  | *Calibrated* | 14.6 (4.3, 24.9) | 2.11 (1.20, 3.69) | 14.0 (2.5, 25.4) | 1.99 (1.11, 3.56) | 1.6 (-11.3, 14.6) | 1.08 (0.58, 2.02) |

Note: High education, high income, and employed groups were used as reference categories. Estimates from imputed datasets were combined using Rubin’s rules. CI: confidence intervals; PD: prevalence difference; PR: prevalence ratio; ; SII: slope index of inequality; RII: relative index of inequality

### **Box S1**. Examples of applying the calibration equations to individual data

We illustrate the application of the calibration equations for height and weight using four example individuals. For each individual, calibrated height (H) and weight (W) are obtained by inserting self‑reported values and covariates into the sex‑specific regression models:

$$H_{\text{calibrated}}=\beta_{0}^{\left( H \right)}+\beta_{1}^{\left( H \right)}\times H_{\text{self}}+\beta_{2}^{\left( H \right)}\times\text{Age}+\gamma_{r}^{\left( H \right)}\times I\left( \text{Region}_{r} \right)+\delta_{e}^{\left( H \right)}\times I\left( \text{Edu}_{e} \right),$$

$$W_{\text{calibrated}}=\beta_{0}^{\left( W \right)}+\beta_{1}^{\left( W \right)}\times W_{\text{self}}+\beta_{2}^{\left( W \right)}\times\text{Age}+\gamma_{r}^{\left( W \right)}\times I\left( \text{Region}_{r} \right)+\delta_{e}^{\left( W \right)}\times I\left( \text{Edu}_{e} \right),$$

where $I\left( \cdot\right)$denotes indicator variables (1 if the category is present, 0 otherwise), (1 if the category is present, 0 otherwise), with high education (Edu) and North region as reference categories. Coefficients are taken from Supplementary Table S6. For simplicity, we assume that self‑reported and measured values are in the same units as in the calibration models (height in cm, weight in kg).

**Example 1.**

Sex: Woman
Self‑reported weight: 65 kg
Age: 30 years
Region: North (reference, all region indicators = 0)
Education: low level (LowEdu = 1, IntermEdu = 0)

Weight calibration (women):

$$\begin{matrix} W_{\text{calibrated}} & =-1.597+1.013\times65+0.043\times30+0\text{ }-0.949\times1\text{ }\left( \text{low education} \right) \\ & =-1.597+65.845+1.29-0.949 \\ & \approx64.6\text{ kg}. \end{matrix}$$

**Example 2.**

Sex: Man
Self‑reported weight: 80 kg
Age: 50 years
Region: Algarve (Algarve = 1, other regions = 0)
Education: intermediate (IntermEdu = 1, LowEdu = 0)

Weight calibration (men):

$$\begin{matrix} W_{\text{calibrated}} & =-1.597+1.013\times80+0.043\times50+0.141 \times1 \left( Algarve \right)-0.512\times1\text{ }\left( \text{intermediate education} \right) \\ & =-1.597+81.04+2.15+0.141-0.512 \\ & \approx81.2\text{ kg}. \end{matrix}$$

**Example 3.**
Sex: Woman
Self‑reported height: 165 cm
Age: 40 years
Region: Centre (Centre = 1, others = 0)
Education: high (highEdu = 1, LowEdu = 0, IntermEdu = 0)

$$\begin{matrix} H_{\text{calibrated}} & =24.678+0.848\times165-0.053\times40+0.728 \times1\text{ }\left( \text{Center} \right) \\ & =24.678+139.92-2.12+0.728 \\ & \approx163.2 \mathrm{cm} \end{matrix}$$

**Example 4.**
Sex: Man
Self‑reported height: 180 cm
Age: 20 years
Region: Madeira (Madeira = 1, others = 0)
Education: low (LowEdu = 1, IntermEdu = 0)

$$\begin{matrix} H_{\text{calibrated}} & =16.342+0.910\times180-0.035\times20-0.026\times1 \left( Madeira \right)-0.153 \\ & =16.342+163.8-0.7-0.026-0.153 \\ & \approx179.3\text{ cm} \end{matrix}$$
